# Sivelestat Sodium in the Treatment of Patients with Acute Respiratory Distress Syndrome Combined with Systemic Inflammatory Response Syndrome

**DOI:** 10.1101/2024.07.28.24311151

**Authors:** Hongli He, Xiaobo Huang

**Affiliations:** Department of Surgical Critical Care, Sichuan People’s Hospital, Chengdu 610072, China

**Keywords:** acute respiratory distress syndrome, systemic inflammatory response syndrome, neutrophil elastase, sivelestat sodium, oxygenation

## Abstract

**Objectives:** Neutrophil elastase (NE) plays an important role in the pathogenesis of acute respiratory distress syndrome (ARDS). Sivelestat sodium, an NE inhibitor, has been approved in Japan for the treatment of patients with ARDS combined with systemic inflammatory response syndrome (SIRS). This trial was designed to evaluate the role of sivelestat sodium in mild-to-moderate ARDS combined with SIRS.

**Methods:** We conducted a multicentre, double-blind, randomized, placebo-controlled trial enrolling patients diagnosed with mild-to-moderate ARDS combined with SIRS admitted within 72 hours of ARDS onset (clinicaltrials.gov, NCT04909697). Patients were randomized in a 1:1 fashion to sivelestat or placebo. Trial drugs were administrated as a 24-hour continuous intravenous infusion at a rate of 0.2 mg/kg/h for 5 days. The primary outcome was PaO_2_/FiO_2_ ratio change on day 3 after randomization, which was defined as (PaO_2_/FiO_2_ ratio on day 3 – baseline PaO_2_/FiO_2_ ratio)/baseline PaO_2_/FiO_2_ ratio.

**Results:** The study was stopped early at the recommendation of an independent Data and Safety Monitoring Board, which noted a between-group difference in mortality. A total of 162 patients were randomized, of whom 81 were assigned to receive sivelestat sodium and 81 placebo. On day 3, the PaO_2_/FiO_2_ ratio improved by 36% in the sivelestat group compared to 3% in the placebo group (difference, 0.27; 95% CI, 0.13 to 0.41, *p* < 0.001). In addition, The invasive mechanical ventilation-free days within 28 days was significantly longer in the sivelestat sodium group compared to the placebo group (median 26.9 days vs. 21.0 days, *p* = 0.004). The Kaplan-Meier curves showed a significant reduction in 90-day mortality in patients receiving sivelestat compared to those not receiving sivelestat (hazard ratio, 0.51; 95% CI, 0.26 to 0.99; log-rank *p* = 0.044).

**Conclusion:** In patients with mild-to-moderate ARDS combined with SIRS, sivelestat sodium may improve oxygenation on day3, increase invasive mechanical ventilation-free days, and was associated with improved survival.

## Introduction

Acute lung injury (ALI) and acute respiratory distress syndrome (ARDS), its most severe form, are the primary drivers of intensive care unit (ICU) admission and are associated with a high rate of mortality[1, 2]. During critical illness, patients are often accompanied with systemic inflammatory response syndrome (SIRS), and these patients are also at a higher risk of ALI/ARDS[3]. Despite advances in critical care management and protective mechanical ventilation strategies[4–6], the treatment of ARDS still remains supportive, and no effective pharmacotherapies are available to improve the mortality of ARDS[7].

It is currently believed that the underlying pathogenesis of ARDS is diffuse injury to pulmonary microvascular endothelial cells and alveolar epithelial cells caused by excessive uncontrolled inflammatory response[1, 8]. Large amounts of cytokines are generated in this cascade-amplified inflammatory response, resulting in direct or indirect tissue damage and consequent organ failure, mediated in part by neutrophil activation and the secretion of cytotoxic substances such as neutrophil elastase (NE)[9, 10]. Preclinical research has found that NE levels in alveolar lavage fluid are significantly increased in animal models of endotoxin- and bacteria-induced ARDS[11–13], and clinical studies have also shown that serum NE levels in patients with ARDS are closely related to the disease severity[14]. Therefore, inhibition of NE would be expected to curb the cascade amplification of inflammatory response in ARDS and alleviate lung injury.

Sivelestat sodium, a small-molecule selective NE inhibitor, was discovered in 1990s[15] and has been shown in previous clinical studies to improve lung function, reduce the duration of mechanical ventilation, shorten ICU stay and improve survival in ALI/ARDS patients, making it the only specific treatment for ARDS currently available[16, 17]. However, most of the clinical studies to date have been retrospective observational studies, and the Phase IV clinical trial of the drug was a non-randomized open-label trial, and the experimental and control groups were set up according to the study sites, and differential post-randomization care at each site may affect the results of the trial[18]. Therefore, we conducted a multi-centre, randomized controlled study to evaluate the role of sivelestat sodium in treating patients with mild to moderate ARDS combined with SIRS.

## Materials and methods

### Trial design and oversight

We conducted an investigator-initiated, double-blind, randomized, placebo-controlled trial at eight centres across China. The ethics committee at each participating centre approved the trial protocol. All patients or their surrogates provided written informed consent. Details of the rationale and design of the study was available in the supplemental materials. Shanghai Huilun (Jiangsu) Pharmaceutical Co., Ltd. supplied the trial drugs but had no role in designing or conducting the trial, or analysing the data and did not have access to the data before publication. We registered the trial at ClinicalTrials.gov (NCT04909697) before recruitment began. An independent Data and Safety Monitoring Committee (DSMB) reviewed the data and performed prespecified blinded interim analyses after the enrollment of 50% of the planned number of participants. This analysis would have led to a recommendation to stop the trial if concerns about participant safety had been raised.

### Study population

All patients who were diagnosed with ARDS were screened for eligibility. Patients were eligible if they were between 18 and 75 years of age, had a PaO_2_/FiO_2_ between 150-300 mmHg, met the criteria for SIRS, and were admitted to one of the participating centres within 72 hours of ARDS onset. The main exclusion criteria were pregnant or lactating women, diagnosed with neutropenia, receiving immunosuppressive agents or high-dose corticosteroid therapy (>40 mg/day), with a known history of severe cardiovascular, respiratory, renal or hepatic disease, or not expected to survive ICU or hospital discharge in the judgement of the attending clinician. Detailed inclusion and exclusion criteria were provided in the Supplementary materials.

### Randomization, blinding and interventions

Randomization was performed by means of a web-based system with the use of computer-generated, permuted-block sequences with stratification according to site. Eligible patients were randomly assigned to receive sivelestat sodium or placebo in a 1:1 ratio. Blinded medication packs were used to ensure allocation concealment. Patients, treating physicians, investigators, data collectors and outcome assessors were unware of the group assignments.

Patients in the sivelestat sodium group received a blinded 24-hour continuous intravenous infusion of sivelastat sodium at a rate of 0.2 mg/kg/h for 5 days after randomization. Patients in the placebo group received a blinded continuous infusion of normal saline at the same rate and according to the same protocol. All other treatments were administered at the discretion of the treating physicians.

### Trial outcomes

The primary outcome was PaO_2_/FiO_2_ ratio change on day 3 after randomization, which was defined as (PaO_2_/FiO_2_ ratio on day 3 – baseline PaO_2_/FiO_2_ ratio)/baseline PaO_2_/FiO_2_ ratio. Secondary outcomes included PaO_2_/FiO_2_ ratio change on day 1 and day 5, PaO_2_/FiO_2_ ratio on day1, 3, 5, mechanical ventilation free days within 28 days, invasive mechanical ventilation free days within 28 days, new-requirement for mechanical ventilation within 28 days, duration of invasive mechanical ventilation, duration of non-invasive mechanical ventilation, duration of high-flow oxygen therapy, duration of nasal catheter oxygen inhalation, duration of mask oxygen inhalation, development of severe ARDS, ICU and hospital-free days within 28 days, 28-day, 60-day and 90-day mortality, incidence of acquired infections during ICU and plasma interleukin-6 (IL-6), NE, C-reactive protein (CRP), IL-10 concerntrations, white blood cell, neutrophil, lymphocyte and platelet count and serum procalcitonin levels, blood CD3+ cell count, CD84 cell count and CD8+ cell count at baseline, day 1, day 3 and day 5.

### Sample size estimation

On the basis of previous studies[19, 20], we hypothesised that the PaO_2_/FiO_2_ ratio change on day 3 after randomization would be 20% in patients with ARDS receiving standard of care. To demonstrate a 14% absolute change in the primary outcome (34% in patients receiving sivelestat sodium) with 80% power at a two-sided alpha level of 0.05, we projected an estimated sample size of 312 participants. The sample size calculation, involving a group sequential design, was performed following the approach of O’Brien and Fleming. According to this method, this trial can be finished as soon as the null hypothesis is rejected in the interim data analysis while controlling the total alpha at the level of 0.05. The interim analyses will take place after the enrolment of 50% patients. Meanwhile, we increase the calculated sample allowing for a potential approximately 4% withdrawal. Finally, a total of 324 patients (162 per group) are needed. Early stopping rule in the interim data analysis (when 162 patients have been enrolled) is based on the O’ Brien-Fleming member of the family of Lan-DeMets spending function rules. The calculation was implemented using the PASS 11.0 software (PASS, NCSS software, Kaysville, USA).

### Statistical analyses

Analyses were conducted using R 4.4.1 software. Two-sided p values < 0.05 were considered statistically significant. Continuous data were reported as means and standard deviations (SD) when normally distributed or as medians and interquartile ranges (IQR) when not normally distributed. The normality of continuous variables will be tested by checking the quantile–quantile (Q–Q) plot. Categorical data will be expressed as numbers and percentages.

We used generalized linear model (GLM) (family = gaussian (link = identity)) to compare the difference in the PaO_2_/FiO_2_ ratio change on day 3 after randomization between groups. For the secondary outcomes, GLM (family = gaussian or binomial (link = identity)) models were used for the continuous or categorical data, respectively. Risk differences and its 95% confidence interval (CI) were calculated for categorical outcomes, and mean differences (95% CI) for continuous outcomes. When the assumptions of GLM models were not fulfilled, log data conversion will be made. Kaplan–Meier curves were used to compare the 28-day survival curves after randomization. The difference between two-groups was calculated by tested by log-rank test. Cox proportional hazards models were performed to calculate the hazard ratios (HR) and associated 95% CI. We tested the assumptions of proportional hazard by checking the plots of Schoenfeld residuals over time.

## Results

### Recruitment and baseline characteristics

From February 2022 through February 2024, we screened 694 patients for eligibility. Of these, 162 patients were enrolled in the trial from six centres across China. At the interim analysis, a between-group difference in mortality was observed and the DSMB stopped recruitment and requested to unblind the data. After reviewing the unblinded data, the DSMB concluded that the trial should be stopped due to the potential mortality benefit of the trial intervention, and the trial was formally stopped. Thus, 81 patients were randomly assigned to the sivelastat sodium group and 81 to the placebo group (**Figure 1**). The numbers of cases from each site were shown in Supplemental Table 1.

**Figure 1.**
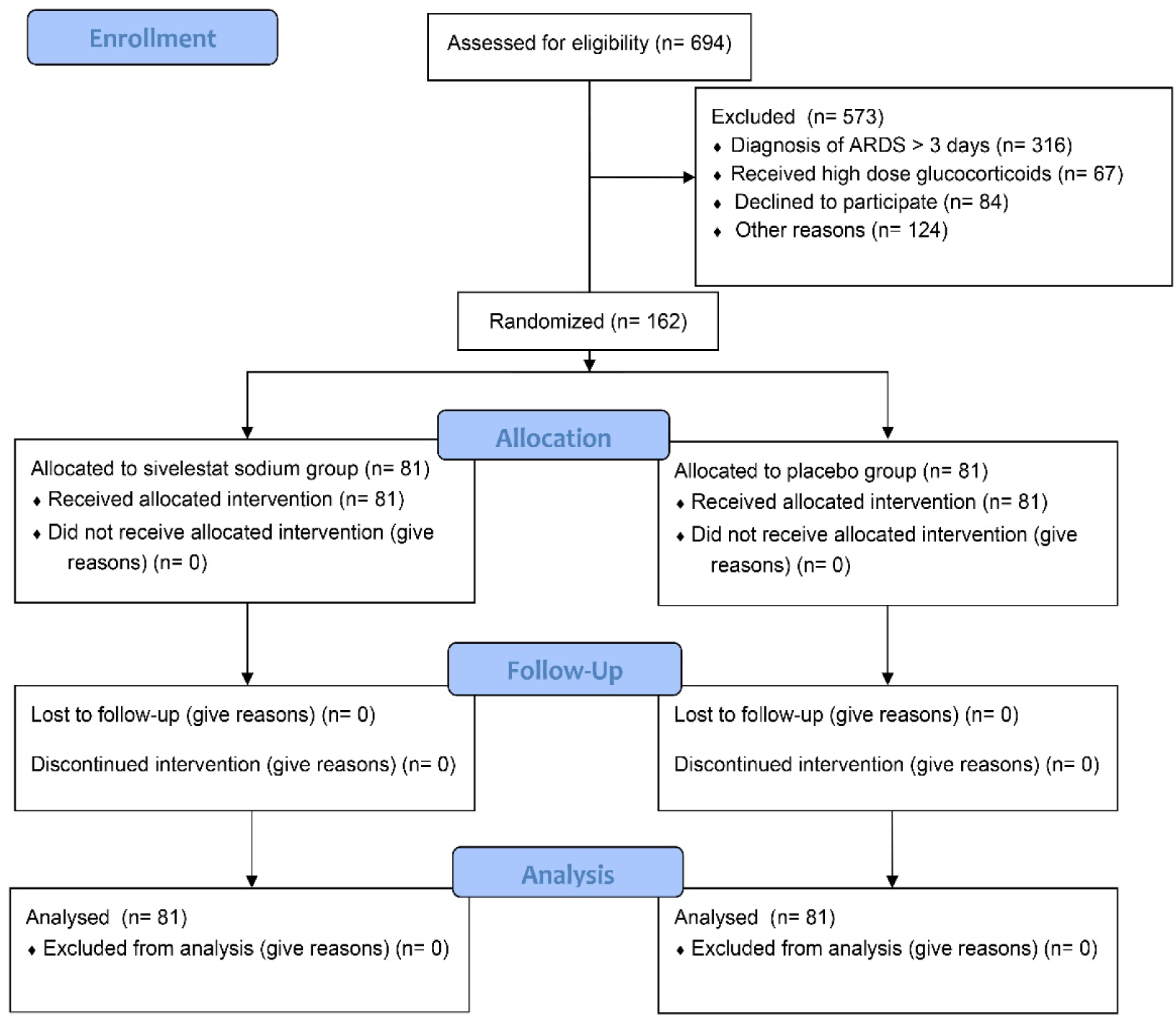
Study flowchart.

Baseline demographics and characteristics were not significantly different between the sivelastat sodium and placebo group (**Table 1**). The median age was 57 years, and 76.5% of patients were men. The majority of patients were receiving mechanical ventilation at enrollment (132/162, 81.5%). The median (IQR) PaO_2_/FiO_2_ ratio was 187.9 (168.8-213.0) mmHg in the sivelastat sodium group and 174.0 (160.0-212.3) mmHg in the placebo group.

**Table 1.** Baseline characteristics of the study subjects.

| Characteristics | Sivelestat Sodium<br>(N=81) | Placebo<br>(N=81) |
| --- | --- | --- |
| Median Age (IQR), yr | 58.0 (45.0-68.5) | 57.0 (48.0-64.5) |
| Gender, n (%) |  |  |
| Women | 19 (23.5) | 19 (23.5) |
| Men | 62 (76.5) | 62 (76.5) |
| Median BMI (IQR), kg/m<sup>2</sup> | 22.3 (21.0-23.5) | 22.6 (21.1-23.0) |
| Comorbidities, no. (%) |  |  |
| Hypertension | 24 (29.6) | 31 (38.3) |
| Diabetes mellitus | 24 (29.6) | 19 (23.5) |
| Chronic kidney disease | 4 (4.9) | 5 (6.2) |
| Cirrhosis | 2 (2.5) | 2 (2.5) |
| Chronic heart disease | 8 (9.9) | 3 (3.7) |
| Cerebrovascular disease | 2 (2.5) | 4 (4.9) |
| Rheumatic disease | 3 (3.7) | 1 (1.2) |
| Tumor | 13 (16.0) | 13 (16.0) |
| Resource of ARDS admission, n (%) |  |  |
| Surgical ward | 6 (7.4) | 6 (7.4) |
| Postoperative care | 24 (29.6) | 16 (19.8) |
| Medical ward | 27 (33.3) | 19 (23.5) |
| Emergency ward or transferred from<br>external hospital | 24 (29.6) | 40 (49.4) |
| Reason for ARDS, n (%) |  |  |
| Pulmonary infection | 32 (39.5) | 38 (46.9) |
| Extrapulmonary infection | 7 (8.6) | 9 (11.1) |
| Pulmonary contusion | 3 (3.7) | 9 (11.1) |
| Burn | 1 (1.2) | 0 (0) |
| Shock | 2 (2.5) | 1 (1.2) |
| Postoperative | 15 (18.5) | 9 (11.1) |
| Acute pancreatitis | 18 (22.2) | 15 (18.5) |
| Others | 3 (3.7) | 0 (0) |
| Use of mechanical ventilation, n (%) | 63 (77.8) | 69 (85.2) |
| Use of glucocorticosteroid, n (%) | 16 (19.8) | 20 (24.7) |
| Clinical parameters |  |  |
| Mean APACHE II score±SD | 12.3±5.2 | 12.0±5.4 |
| Median SOFA score (IQR) | 4 (3-7) | 5 (4-6) |
| Median number of organ failure (IQR) | 2 (2, 3) | 2 (2, 3) |
| Median lac (IQR), mmol/L | 1.7 (1.1-2.4) | 1.7 (1.3-2.3) |
| Median PaO<sub>2</sub>/FiO<sub>2</sub> ratio (IQR) | 187.9 (168.8-213.0) | 174.0 (160.0-212.3) |
| Laboratory parameters, median (IQR) |  |  |
| Serum CRP, pg/mL | 3379.6 (2393.7-4533.0) | 3290.4 (2508.6-4495.1) |
| Serum IL-6, pg/mL | 72.5 (32.7-189.2) | 86.3 (49.4-172.3) |
| Serum IL-10, pg/ml | 15.3 (7.2-32.4) | 15.8 (6.8-31.1) |
| Neutrophil elastase, pg/ml | 506.3 (392.1-872.5) | 567.9 (309.5-1183.9) |
| Respiratory parameters |  |  |
| Median FiO<sub>2</sub> (IQR) | 0.45 (0.40, 0.50) | 0.50 (0.40, 0.60) |
| Median PEEP (IQR), mmHg | 9.0 (5.0, 10.0) | 8.0 (7.0, 10.0) |

IQR denotes interquartile range, BMI denotes body mass index, ARDS denotes acute respiratory distress syndrome, APACHE II denotes Acute Physiology and Chronic Health Evaluation II, SD denotes Standard Deviation, SOFA denotes sequential organ failure assessment, CRP denotes C-reactive protein, IL-6 denotes interleukin-6, IL-10 denotes interleukin-10.

### Primary and secondary outcomes

On day 3 after randomization, the PaO_2_/FiO_2_ ratio improved by 36% in the sivelestat sodium group compared to 3% in the placebo group (difference, 0.27; 95% CI, 0.13 to 0.41, *p* < 0.001). The results remained significant after adjustment for study sites (*p* = 0.003). The results were similar on day1 and day5 (**Table 2**).

**Table 2.**
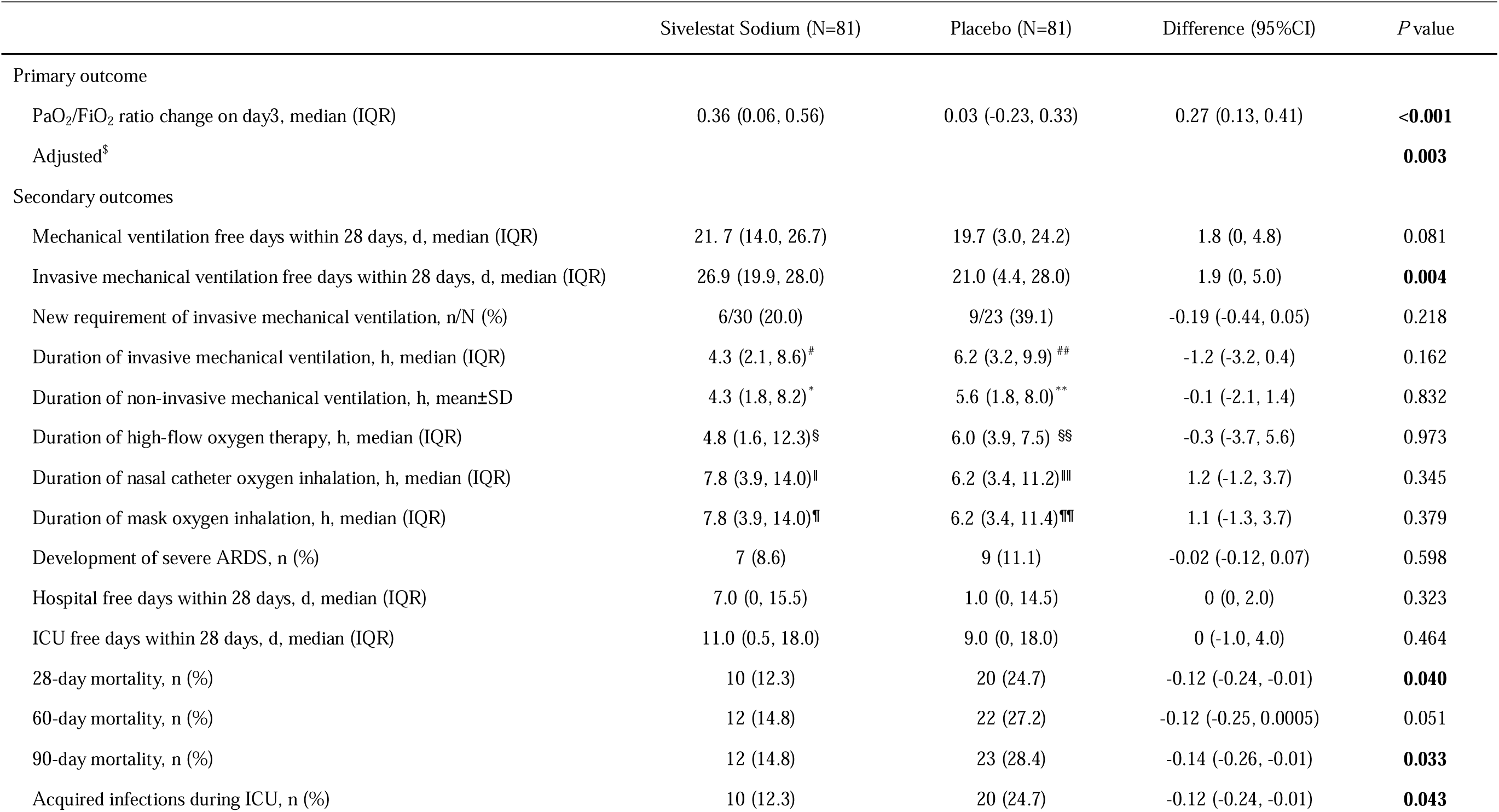
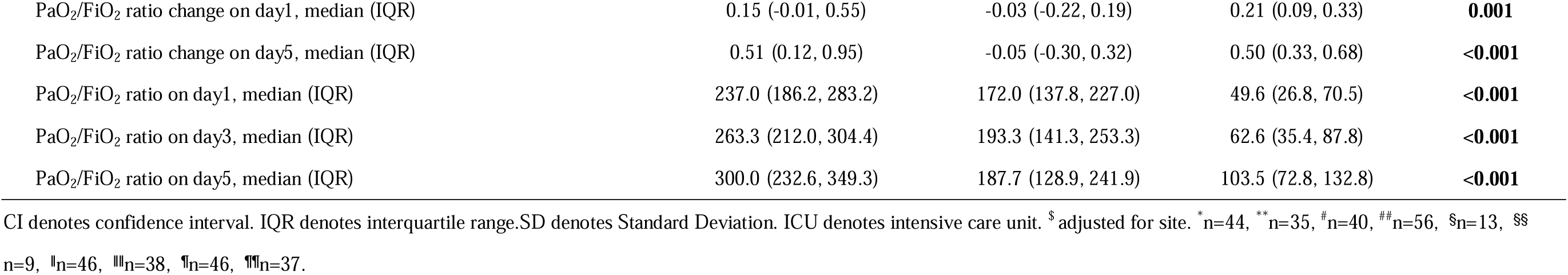
Primary and main secondary endpoints.

The invasive mechanical ventilation free days within 28 days was significantly longer in the sivelestat sodium group compared to the placebo group (median 26.9 days vs. 21.0 days, *p* = 0.004). Ten patients (12.3%) in the sivelestat sodium group and 20 patients (24.7%) in the placebo group developed acquired infections during the ICU stay (*p* = 0.043). The 28-day, 60-day and 90-day mortality were significantly different between patients with and without sivelestat administration (12.3% vs. 24.7%, p = 0.040; 14.8% vs. 27.2%, p = 0.051; 14.8% vs. 28.4%, p = 0.033, respectively). The Kaplan-Meier curves showed a significant reduction in 90-day mortality in patients treated with sivelestat sodium compared with those not treated (hazard ratio, 0.51; 95% CI, 0.26 to 0.99; log-rank *p* = 0.044) (**Figure 2**).

**Figure 2.**
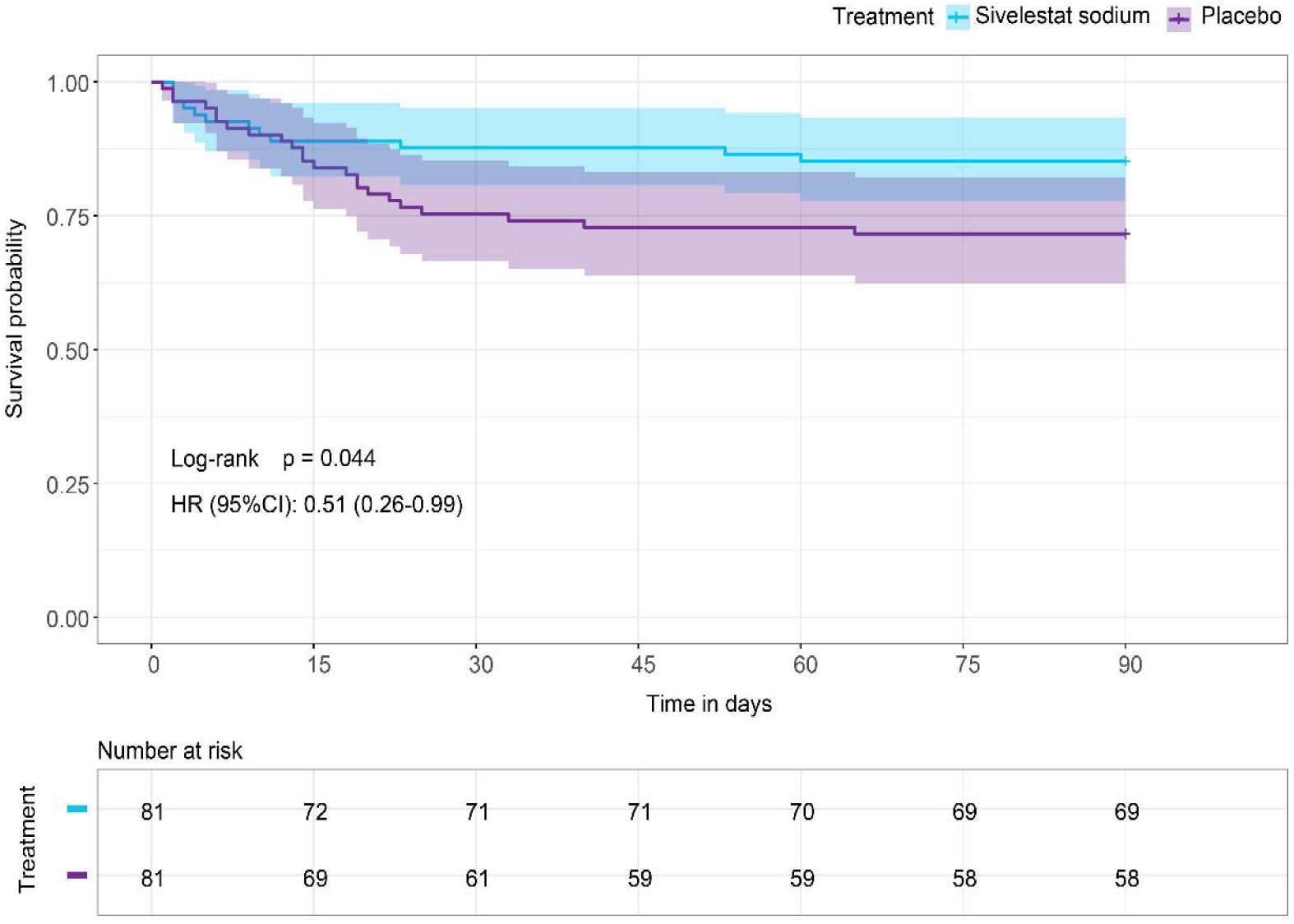
Survival curve. HR denotes hazard ratio. CI denotes confidence interval.

There were no significant differences between the two groups in new-requirement of mechanical ventilation, the duration of non-invasive mechanical ventilation, and the duration of other oxygen therapies, including high-flow nasal oxygen (HFNO), nasal catheter oxygen inhalation and mask oxygen inhalation. The ICU free days within 28 days and hospital-free days within 28 days were both comparable between two groups (**Table 2**).

For secondary laboratory outcomes, no difference was observed between groups for serum NE, IL-6, CRP and IL-10 levels (**Figure 3**), white blood cell, neutrophil, lymphocyte and platelet counts (Supplementary figure 1), as well as serum procalcitonin levels, blood CD3+ cell count, CD84 cell count and CD8+ cell count (Supplementary figure 2) at all study time points (day 1, day 3 and day 5).

**Figure 3.**
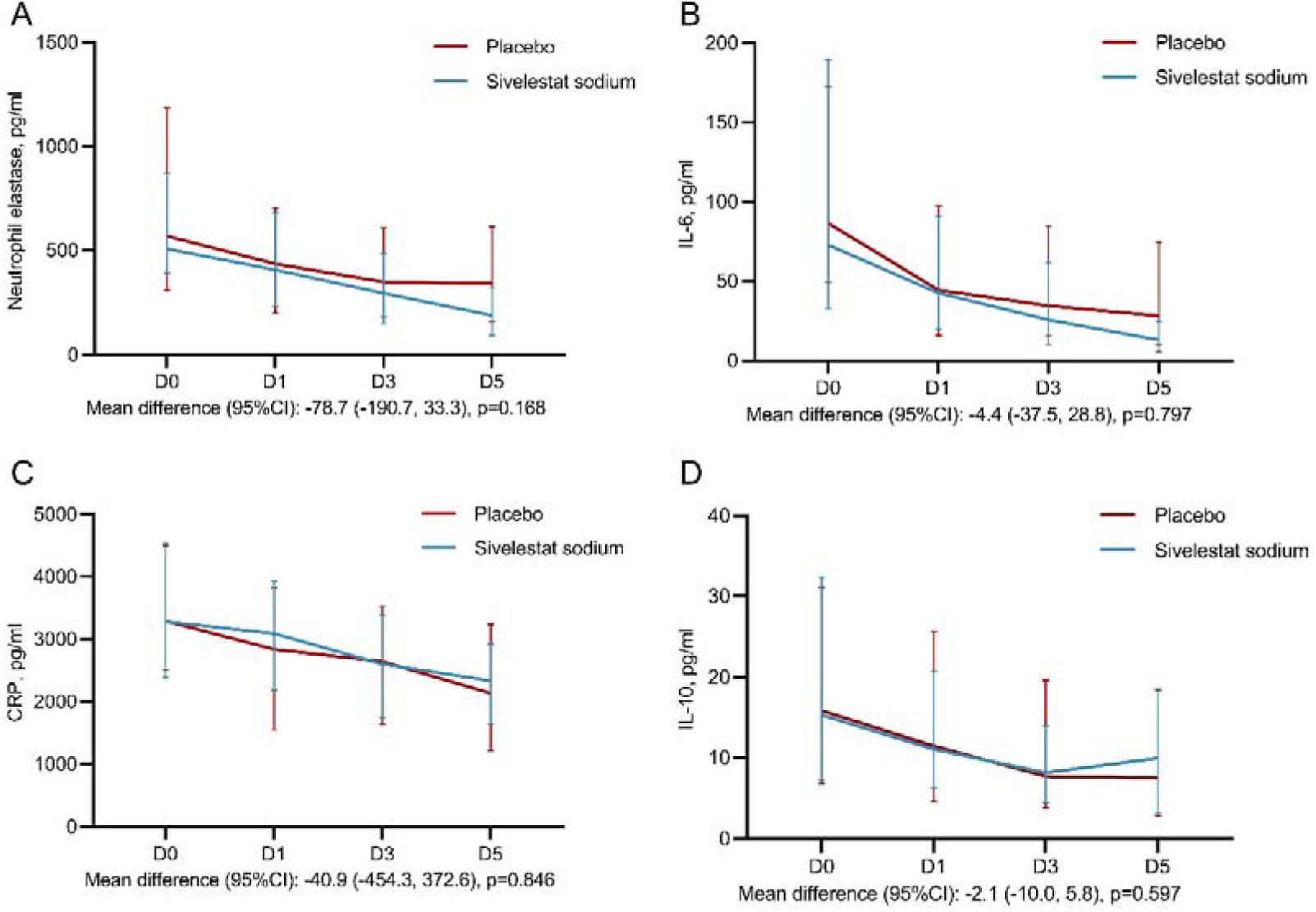
Laboratory indexes. CRP denotes C-reactive protein, IL-6 denotes interleukin-6, IL-10 denotes interleukin-10, CI denotes confidence interval.

### Adverse events

We observed no significant differences between the two groups in the number of patients with prespecified adverse events. Details regarding adverse events are provided in Supplementary Table 2.

## Disscussion

In this multicentre, double-blind, randomized, placebo-controlled trial, we compared the continuous infusion of sivelestat sodium with placebo (normal saline) for improvement in oxygenation on day 3 after randomization in adult patients with mild-to-moderate ARDS combined with SIRS. Sivelestat sodium infusion resulted in a significantly increased PaO_2_/FiO_2_ ratio on day 3 than placebo. Moreover, patients who received sivelestat sodium had longer invasive mechanical ventilation-free days and a lower incidence of acquired infections during the ICU stay compared to patients who did not receive sivelestat sodium. Also, the 28-day, 60-day and 90-day mortality were significantly lower in the sivelestat sodium group than in the control group.

In 2002 Japan was the first country to approve sivelestat for the treatment of “ALI associated with SIRS”. After the approval, a post-marketing clinical study was conducted at the request of the Pharmaceuticals and Medical Devices Agency to re-evaluate the safety and efficacy of sivelestat in actual clinical settings in Japan. This phase IV study included 404 sivelestat-treated patients and 177 controls. The sivelestat group showed a significant improvement in the primary endpoint of ventilator-free days (VFDs) compared to the control group[18]. In addition, the adjusted 180-day survival rate was significantly higher in the sivelestat group than in the control group. On the other hand, the STRIVE study, conducted in six countries other than Japan, randomized 492 mechanically ventilated patients with ALI/ARDS and failed to demonstrate the efficacy of sivelestat[21]. Furthermore, an increase in 180-day all-cause mortality was noted in the sivelestat group and the study was halted on the recommendation of the DSMB.

The discrepant results between the STRIVE study and the Japanese phase IV study may be due to differences in patient population and study design. The Japanese study was an open-label, non-randomized clinical trial, and the experimental and control groups were set up according to the study centres, and the differences between the centres in the treatment of ALI/ARDS may have influenced the study results. Patients enrolled in the Japanese study had less severe respiratory function and less organ dysfunction than patients in the STRIVE study, and patients with burns or trauma were excluded. In addition, the Japanese study defined SIRS as an inclusion criterion, which helps to identify ARDS patients with a pro-inflammatory phenotype[22], whereas the STRIVE study did not. A post-hoc analysis of the STRIVE patients involving those who had a mean Lung Injury Score less than 2.5, showed favourable trends in mortality and VFDs in patients receiving sivelestat[23]. Therefore, it is important to select appropriate patients to be treated with sivelestat. The results of these two trials suggest that sivelestat may be effective in patients with relatively mild ARDS and with a typical pro-inflammatory phenotype, which were the inclusion criteria for our study.

Since NE may also have a beneficial bactericidal effect[24], inhibiting NE may increase the risk of infection. A preclinical study investigating the effect of sivelestat in an animal model of *S. pneumoniae*-induced lung injury showed that sivelestat could reduce the bacterial counts in bronchoalveolar lavage fluid (BALF) and lung interstitial tissue and preserve the host immune response[13]. Clinical data further supported this finding that sivelestat did not worsen infection. In both the STRIVE study and the Japanese Phase IV study, there was no significant difference in the incidence of serious adverse events related to infection between the sivelestat group and the control group. In addition, our study suggests that sivelestat may reduce the incidence of acquired infections during ICU stay.

The study had several limitations. First, the current sample size was not powered to detect a difference in mortality, and the stopping of the trial midway further weakens the robustness of the results. Therefore, the results of this study should be interpreted with caution. Second, subjective factors contribute to the decision to wean patients from mechanical ventilation, which may bias the results of the duration of mechanical ventilation. Finally, we did not observe the effects of sivelestat on long-term outcomes, such as 180-day mortality.

## Conclusion

In this trial involving adult patients with mild-to-moderate ARDS combined with SIRS, the infusion of sivelestat sodium significantly improved oxygenation on day 3 after randomization, increased invasive mechanical ventilation-free days, and was associated with improved survival. Further large-scale RCTs are warranted to confirm the effects of sivelestat sodium on mortality in patients with ARDS combined with SIRS.

## Data Availability

All data produced in the present study are available upon reasonable request to the authors. And all data produced in the present work are contained in the manuscript.

## Date availability

All data produced in the present work are contained in the manuscript.

## Ethics and approval statement

The protocol and the informed consent document have been reviewed and approved by the Institutional Ethics Committee of all participating centers. Study investigators will provide potential participants with verbal and written information prior to inclusion in the study. Informed consent will be provided from participants or their authorized representatives. The study was registered in the ClinicalTrials.gov (NCT04909697).

## Authorship contribution statement

Hongli He: Patient screening, data collection and analysis and interpretation of data; Xiaobo Huang: conceived, designed and supervised the study; analysis and interpretation of data; critical revision of the manuscript; obtained funding. All authors have read the manuscript and approved its submission.

---

**Figure S1.**
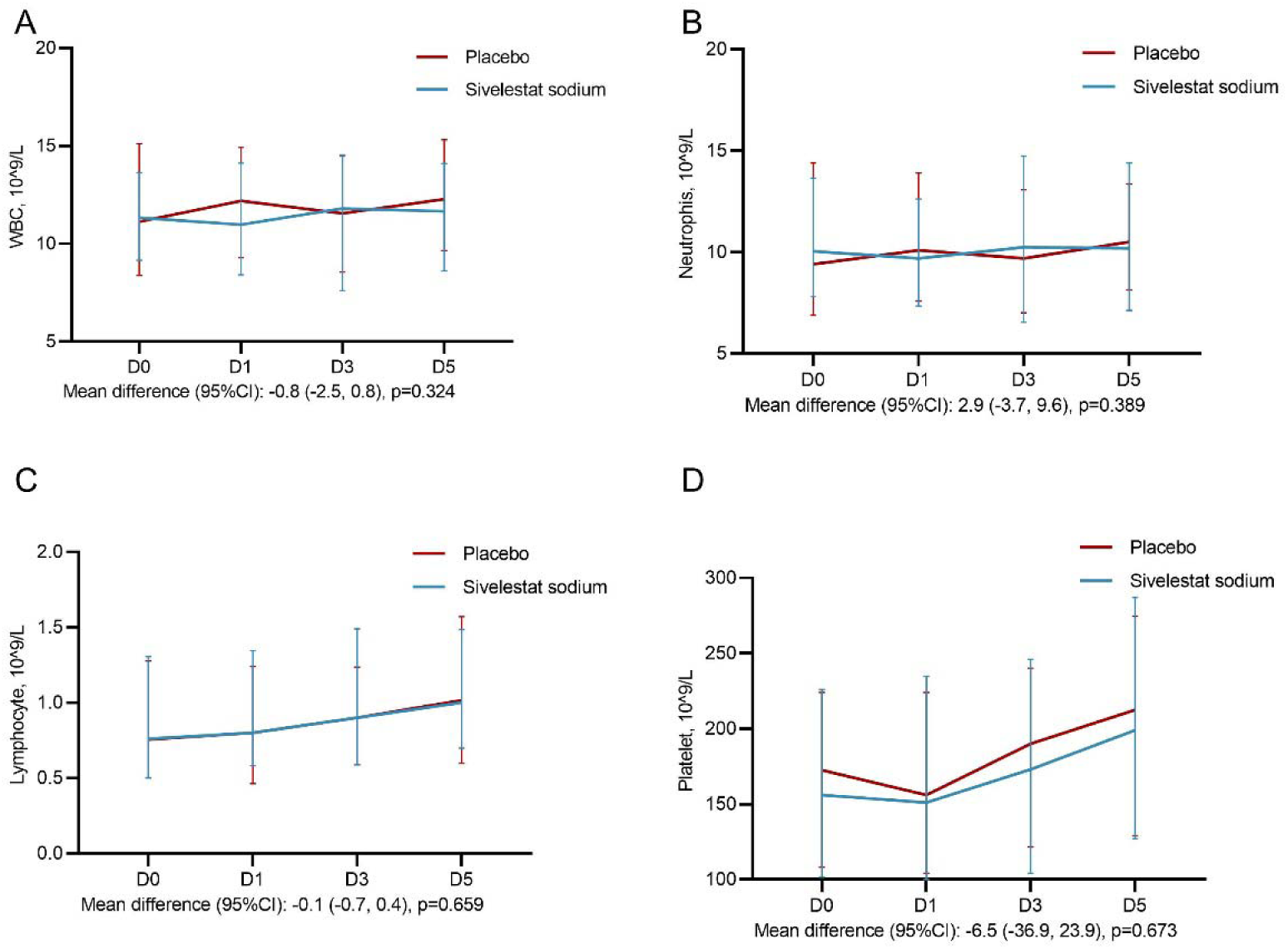
Laboratory indexes.

**Figure S2.**
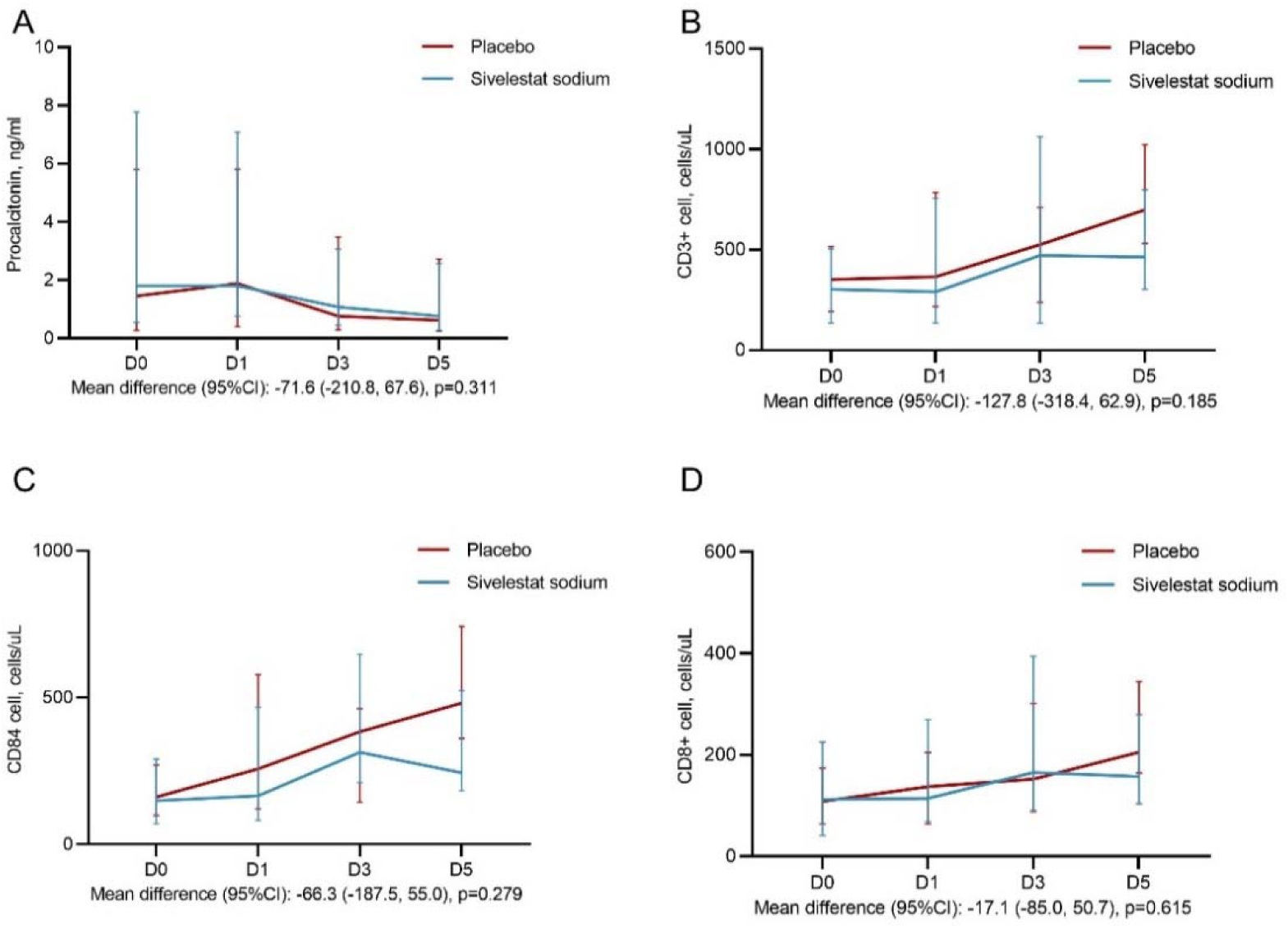
Laboratory indexes.

**Table S1.** Numbers of cases from each site.

| Participating sites | Total | Sivelestat Sodium | Placebo |
| --- | --- | --- | --- |
| Sichuan Provincial People's Hospital | 68 | 35 | 33 |
| Affiliated Hospital of Chengdu University of Traditional Chinese Medicine | 5 | 2 | 3 |
| Affiliated Hospital of Chengdu University | 2 | 2 | 0 |
| Chengdu Fifth People's Hospital | 31 | 14 | 17 |
| Chengdu Shuangliu District First People's Hospital | 27 | 12 | 15 |
| Chengdu Second People's Hospital | 29 | 16 | 13 |
| Total cases | 162 | 81 | 81 |

**Table S2.** Adverse events.

| Adverse event | Sivelestat Sodium (N=81) | Placebo (N=81) |
| --- | --- | --- |
| Total no. of adverse events | 9 | 13 |
| Acute liver injury | 4 | 7 |
| Dyspnea | 1 | 2 |
| Leukopenia | 2 | 3 |
| Essential thrombocythemia | 1 | 1 |
| Elevated creatinine/urea | 1 | 0 |

## Ethical apprval

| Organization | Ethical state | Number (no.) | Date |
| --- | --- | --- | --- |
| Sichuan People's Hospital | Approve | 2022-02-07 | 2022-02-07 |
| Affiliated Hospital of Chengdu University of Traditional Chinese Medicine | Approve | 2022-07-14 | 2022-07-14 |
| Chengdu Shuangliu District first People's Hospital | Approve | 2022-05-12 | 2022-05-12 |
| Chengdu Second People's Hospital | Approve | 2022-06-22 | 2022-06-22 |
| Chengdu Fifth People's Hospital | Approve | 2022-06-02 | 2022-06-02 |
| Affiliated Hospital of Chengdu University | Approve | 2023-08-08 | 2023-08-08 |
| Chengdu Third People's Hospital | Approve | 2022-12-01 | 2022-12-01 |
| Sichuan Cancer Hospital | Approve | 2023-07-04 | 2023-07-04 |

